# Fecal Microbiota Transplantation Donor Screening: Is *Dientamoeba fragilis* a Valid Criterion for Donor Exclusion? A Longitudinal Study of a Swiss Cohort

**DOI:** 10.1101/2025.10.29.25338826

**Authors:** Keyvan Moser, Aurelie Ballif, Trestan Pillonel, Maura Concu, Elena Montenegro Borbolla, Beatrice Nickel, Camille Stampfli, Marie-Therese Ruf, Maxime Audry, Nathalie Kapel, Susanna Gerber, Damien Jacot, Claire Bertelli, Tatiana Galperine

**Affiliations:** Service of Infectious Diseases, Lausanne University Hospital and University of Lausanne, Lausanne, Switzerland; French Group for Fecal Microbiota Transplantation (GFTF), Paris, France; Department of policlinics, Centre for Primary Care and Public Health (Unisanté), Lausanne; Switzerland; Swiss Tropical and Public Health Institute, Allschwil, Switzerland; University of Basel, Basel, Switzerland; Service of Pharmacy, Lausanne University Hospital and University of Lausanne, Lausanne, Switzerland; Laboratoire de Coprologie fonctionnelle, APHP, GH Pitié-Salpêtrière, Paris, France; Université Paris Cité, INSERM S1139, Paris, France; Institute of Microbiology, Lausanne University Hospital and University of Lausanne, Lausanne, Switzerland

**Keywords:** Fecal Microbiota Transplantation, *Dientamoeba fragilis*, *Clostridioides difficile*, Patient Safety, Treatment Efficacy, Shotgun Sequencing, Donor Screening

## Abstract

**Introduction:** *Dientamoeba fragilis* is a protozoan of the human digestive tract, yet its transmission and pathogenic role remain poorly understood. This study aimed to evaluate its impact on the efficacy and safety of FMT in treating recurrent *Clostridioides difficile* infection (rCDI).

**Patients and methods:** This longitudinal cohort study analyzed stool samples from FMT donors and recipients pre-treatment and at 2 and 8 weeks post-FMT. All samples were retrospectively tested using real-time PCR. Shotgun metagenomics was also performed on selected donor-recipient pairs to explore transmission. CDI cure rates, gastrointestinal adverse events (AE), and serious adverse events (SAE) were assessed prospectively.

**Results:** A total of 53 FMT were analysed (179 samples), with 23 (43%) derived from *D. fragilis*-positive donor stool (4 of 10 donors, 40%). Four of 52 recipients (18.2%), initially negative and who received treatment from positive donors, tested positive post-FMT. Shotgun metagenomics could not definitely confirm transmission due to the lack of a good reference genome. No significant differences in efficacy, AE, or SAE were observed between FMT from *D. fragilis*-positive versus -negative donors, even in immunocompromised patients. No SAE were attributed to FMT.

**Conclusion:** *D. fragilis* may be transmitted via FMT without clinical impact, highlighting the need to reconsider donor exclusion and aligning with growing evidence of its questionable pathogenicity.

## INTRODUCTION

Since 2014, fecal microbiota transplantation (FMT) has been the recommended treatment for recurrent *Clostridioides difficile* infections (rCDI), with success rates up to 85% and improved patient survival [1–5]. FMT aims to therapeutically modulate the recipient’s gut microbiome.

For safety reasons, donors undergo rigorous selection procedures, including multiple clinical and biological screenings, to minimize the risk of transmitting pathogens and potential diseases associated with gut microbiota [6]. Following this selection process, only 3-10% of candidates qualify as eligible fecal microbiota donors, limiting the availability and accessibility of this proven beneficial treatment [7] [8].

*Dientamoeba fragilis* is an intestinal protozoan belonging to the *Trichomonadidae* order even though it lacks flagella [9]. *D. fragilis* has been detected worldwide with prevalence rates ranging from 0.2% to 82%, and often exceeding 20% in molecular epidemiological studies [10]. However, its route of transmission remains poorly understood. The pathogenicity of *D. fragilis* remain debated, with limited evidence linking it to gastrointestinal symptoms such as diarrhea and abdominal pain [11].

FMT serves as a valuable and reliable model for studying the impact of controversial fecal microorganisms, providing direct exposure to donor gut microbiota in recipients. For example, *Blastocystis hominis*, another protist with debated pathogenicity, was removed from the donor exclusion list following a study by Teerver et al., which demonstrated no adverse consequences in recipients from *B. hominis*-positive donors [12]. As a result of this study, the 5th edition of the Guide to the Quality and Safety of Organs for Transplantation (EDQM, 2022) no longer lists *B. hominis* as an exclusion criterion for donors, unlike *D. fragilis*. The transmission and impact of *D. fragilis* during FMT, however, still require further investigation.

To date, real-time PCR (RT-PCR) for *D. fragilis* is not part of routine donor screening at the Lausanne University Hospital, the only center in Switzerland performing FMT. The primary objective of our study was to assess the prevalence of *D. fragilis* in stool samples from FMT donors and their paired recipients, both before and after FMT, using RT-PCR. Additionally, we aimed to investigate whether the presence of *D. fragilis* affects the efficacy of FMT or is associated with adverse events post-FMT.

## METHODS

### Study design and setting

This observational cohort study was conducted at Lausanne University Hospital (CHUV) in Switzerland. From January 2023 to December 2024, all eligible patients receiving FMT as part of routine clinical care for rCDI and their corresponding donors involved in the production of FMT batches used for treatment were prospectively included. Molecular screening for *D. fragilis* was retrospectively performed using RT-PCR-based detection methods on stool samples collected from donors and from their paired recipients before and after FMT. The patient sample size was determined based on pragmatic considerations, and inclusion followed a total enumerative sampling approach, encompassing all patients who met the eligibility criteria over a two-year period. Study data were collected from patients’ electronic health records and were documented and managed using REDcap electronic data capture tools hosted at CHUV. This manuscript was prepared in accordance with the STROBE statement for cohort studies. [13].

### Study participants

#### Patients – rCDI

Eligibility criteria required patients to be at least 18 years old and to have undergone FMT, with the indication aligned with the European Society of Clinical Microbiology and Infectious Diseases (ESCMID) guidelines for *Clostridioides difficile* treatment [1].

Patients received at least 10 days of pretreatment with fidaxomicin or vancomycin. FMT was administered to inpatients either via endoscopy (colonoscopy or jejunostomy) following bowel lavage with 2 liters of macrogol solution the day prior or through oral ingestion of 20 capsules per day over two consecutive days (40 in total). In severe cases, the dosage was increased to 30 capsules per day (60 in total).

Patient variables were standardized, including age (continuous variable), sex (female or male), and immunosuppression status (binarized). Severe immunosuppression status was defined as current or foreseeable neutropenia (<⍰1500 neutrophils/µl) within the next 14 days; scheduled or recent (<⍰110 days) allogeneic stem cell transplantation; active graft-versus-host disease (GvHD) requiring immunosuppressive treatment; or ongoing chemotherapy.

To assess the clinical success of FMT, cure was defined as the absence of CDI recurrence at 8 weeks post-FMT. Criteria for severe, complicated and recurrent CDI followed the ESCMID guidelines [1]. Gastrointestinal adverse events (AE) following FMT, including bloating, nausea, abdominal pain, and changes in bowel movements, were prospectively monitored and documented at weeks 2 and 8. Follow-up assessments were conducted via phone at 6 months, 12 months, and annually for up to 5 years.

Serious adverse events (SAE) were evaluated in accordance with the ICH E2A guidelines [14]. The causal relationship between FMT and SAE was evaluated using categories defined in the Fecal Microbiota Transplant National Registry of the American Gastroenterological Association (AGA) [15].

#### Donors

All microbiota donors were healthy, unpaid, volunteers aged 18 to 50 years and were submitted to a detailed biological screening (Supplementary Table 1). Stool donations were collected at the CHUV FMT center and processed individually, without pooling between donations or donors (see Supplementary material for detailed FMT production). The donors included in this study were those who provided stool used in the production of the treatment for the enrolled patients. In all samples, direct microscopic examination for *D. fragilis* yielded negative results.

### Native stool collection, *D. fragilis* detection

Donor native stool used for treatment was collected prior to treatment production. Stool samples from FMT recipients were obtained 24-48 hours before FMT, as well as at 2 and 8 weeks post-FMT, as part of routine care for traceability purposes. All samples were aliquoted and biobanked at -80°C (BIOSTOOL, BB_041). The diagnosis of *D. fragilis* was conducted using two in-house RT-PCRs at the Diagnostic Center of Swiss Tropical and Public Health Institute (Swiss TPH) in Allschwil (see Supplementary Material for detailed detection methods). A cut-off cycle threshold (Ct) of <40 was used for both PCRs. The analysis of shotgun metagenomics sequences of positive paired donor and recipient stools was performed at the Genomics and Metagenomics laboratory of the CHUV (see Supplementary Material for detailed methods).

### Outcomes

The primary outcome was the assessment of the prevalence of *D. fragilis* in stool samples using RT-PCR from both donors and corresponding recipients before FMT, and at weeks 2 and 8 post-FMT. Secondary outcomes included comparing the efficacy, gastrointestinal AE and SAE associated with FMT, depending on the *D. fragilis* status of donor batches.

### Statistics

For the primary objective, the prevalence of *D. fragilis*–positive RT-PCR in donors and recipients were expressed as rates. With regard to the secondary objectives, the analysis focused on identifying differences in CDI recurrence rates and gastrointestinal AE and SAE between recipients of *D. fragilis*-positive and *D. fragilis*-negative treatments, using Chi-square or Fisher’s exact test. A p-value below 0.05 was considered indicative of statistical significance in all comparisons. All analyses were performed using RStudio, version 4.2.1.

### Ethical statement

Ethical approval for this study was obtained from the Canton of Vaud Ethics Committee (ID: CER-VD 2024-00418). Written informed consent was obtained for anonymized patients and donors for participation in the research and reuse of biological samples.

## RESULTS

### RT-PCR detection of *D. fragilis* in stools from donors and FMT recipients

A total of 179 stool samples were analyzed by RT-PCR for *D. fragilis* including 37 samples from 10 donors and 142 samples from 47 recipients. All donors and recipients were of European ethnic origin. The characteristics of the study population are summarized in Table 2. Details of recipients’ characteristics, clinical follow-up, RT-PCR results, and FMT data are provided in Supplementary Table 2.

**Table 1.**
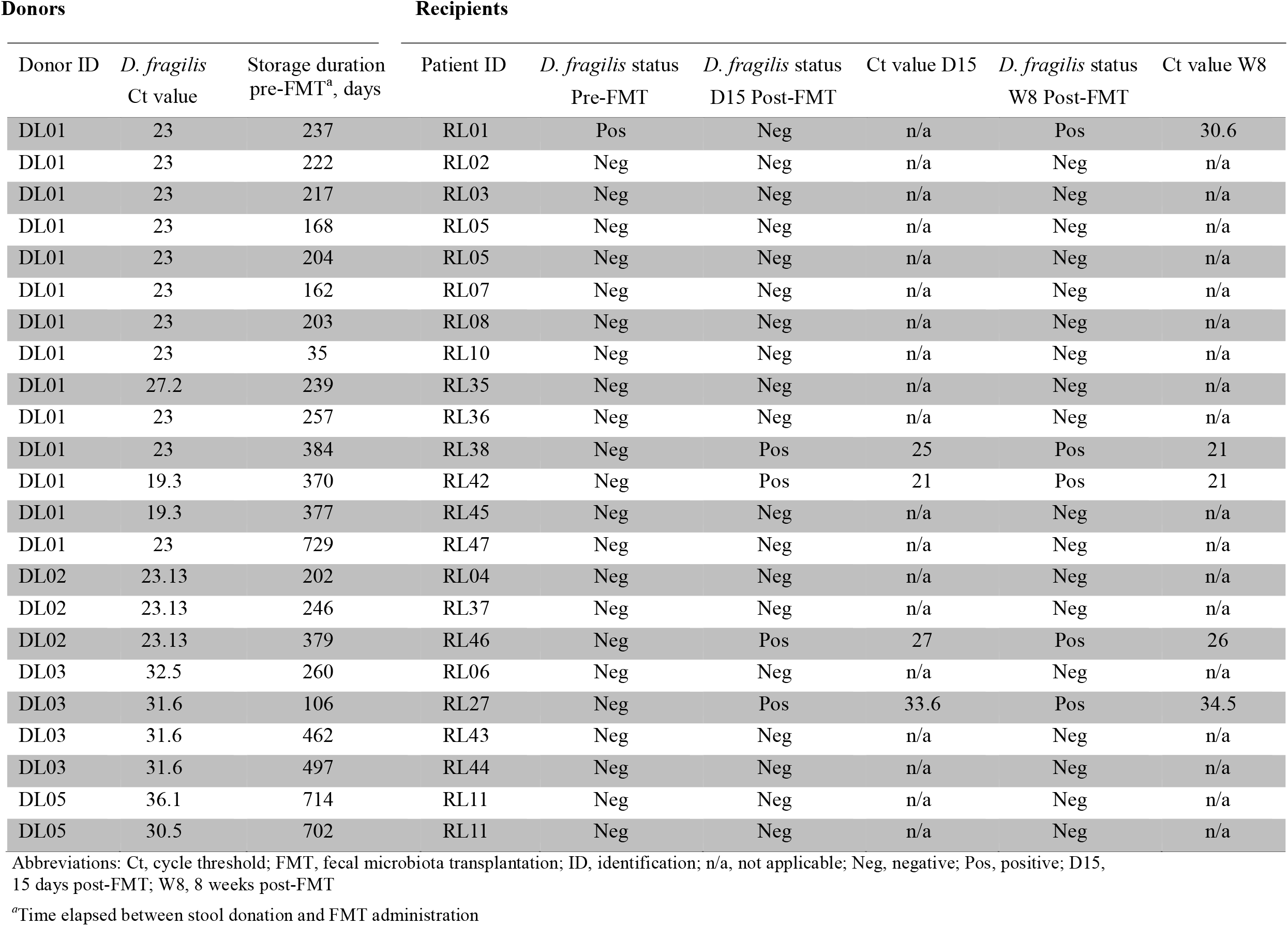
Fecal microbiota transplantation with *D. fragilis*-positive donor stool and recipient *D. fragilis* status.

**Table 2.**
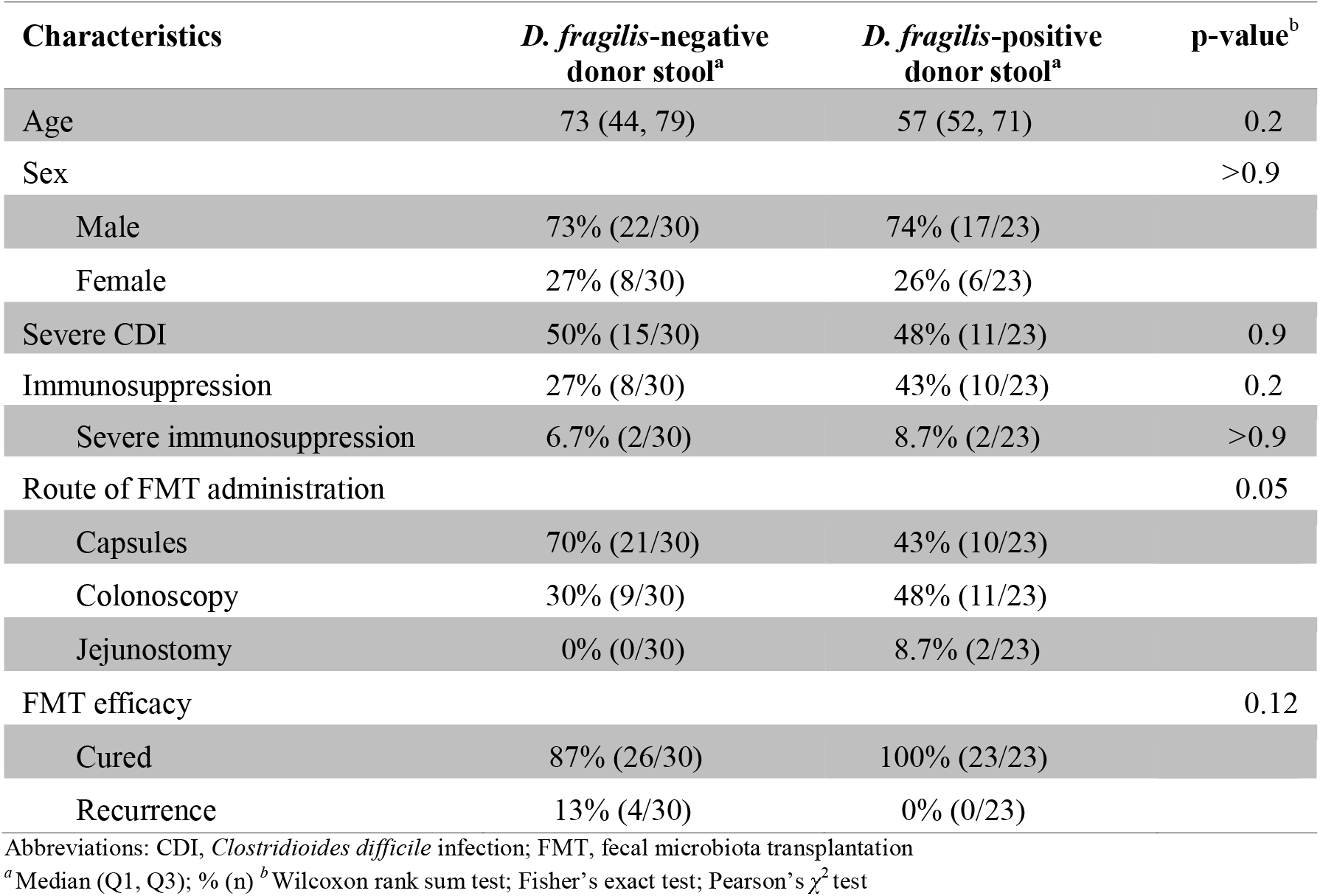
Demographic characteristics of fecal microbiota transplantation recipients and comparison between transfer of *D. fragilis*-positive and -negative donor stool.

*D. fragilis* was detected in the stool of 4 out of 10 donors (40%), with Ct values for RT-PCR ranging from 19.3 to 36.1 (Table 1). Fecal material from the 10 donors was used to produce a total of 233 oral FMT preparations (9,320 capsules) and 54 suspensions. Batches derived from *D. fragilis*-positive donor stool accounted for 46 of the 233 oral preparations (1,840 capsules, 19.7%) and 37 of the 54 suspensions (68.5%). In total, 53 FMT treatments were administered, of which 23 (43%) originated from *D. fragilis*-positive donor stool.

Prior to FMT, *D. fragilis* was negative in 52 of 53 (98%) recipient stool samples. One recipient (1/47, 2%) was tested positive before FMT and received a treatment coming from *D. fragilis*-positive donor stool. There is a significant difference in the prevalence of *D. fragilis* between donors and recipients in our sample (p<0.05). Among the 22 patients who tested negative before FMT and received stool from *D. fragilis*-positive donors, only four (18.2%) tested positive by RT-PCR at day 15 post-FMT, all of whom remained positive at week 8. All four patients received FMT via colonoscopy, with transplants thawed using a standardized warm water bath method (34.5°C). Despite varying immunosuppression statuses, these patients tolerated the procedure well and were considered cured at the 8-week follow-up. The *D. fragilis* PCR Ct values in stool samples from positive donors used for their treatments ranged from 19.3 to 31.6. In positive recipients’ stool, Ct ranged from 21 to 34.5. Notably, four other patients who received FMT from the same donor batches under identical conditions did not test positive for *D. fragilis* at any point during follow-up.

Shotgun metagenomic sequencing was performed on one stool sample from donor DL01 and two samples from the FMT recipient RL42, as this pair presented the highest *D. fragilis* concentration (lowest Ct). From the 36 to 51 million reads generated per sample, only 19 (DL01), 380 (RL42 D15), and 31 (RL42 W8) reads mapped to the available *D. fragilis* transcriptome. Due to the extremely low number of mapped reads, we were unable to confirm with confidence the transmission of *D. fragilis* from donor to recipient.

### Efficacy at 8 weeks according to *D. fragilis* positivity of the donors

At week 8, CDI resolution was achieved in 44/47 (93.6%) patients after a single FMT, increasing to 46/47 (97.9%) after two FMTs and 47/47 (100%) after three. Notably, all post-FMT CDI recurrences occurred in recipients of stool from *D. fragilis*-negative donors. The presence of *D. fragilis* in donor stools had no impact on FMT efficacy (p=0.12) (Table 2). The median follow-up at the time of analysis was 13.3 months (range 3-25). Even though the cohort included individuals with significant morbidity and severe immunosuppression (see Supplementary Table 2), no new episodes of CDI were reported during the studied period after the 8 weeks follow-up.

### AE and SAE at 15 days and 8 weeks post FMT

Patients could experience multiple gastrointestinal AE associated with a single FMT. A total of 53 gastrointestinal AE and 12 SAE were recorded and are detailed in Supplementary Table 2. None of the SAE were related to FMT. Four SAE (33%) occurred in recipients with transplants from *D. fragilis*-positive donors. There were no significant differences in the incidence of gastrointestinal adverse events or severe adverse events between patients treated with FMT from *D. fragilis*-positive or -negative donors (Table 3).

**Table 3.**
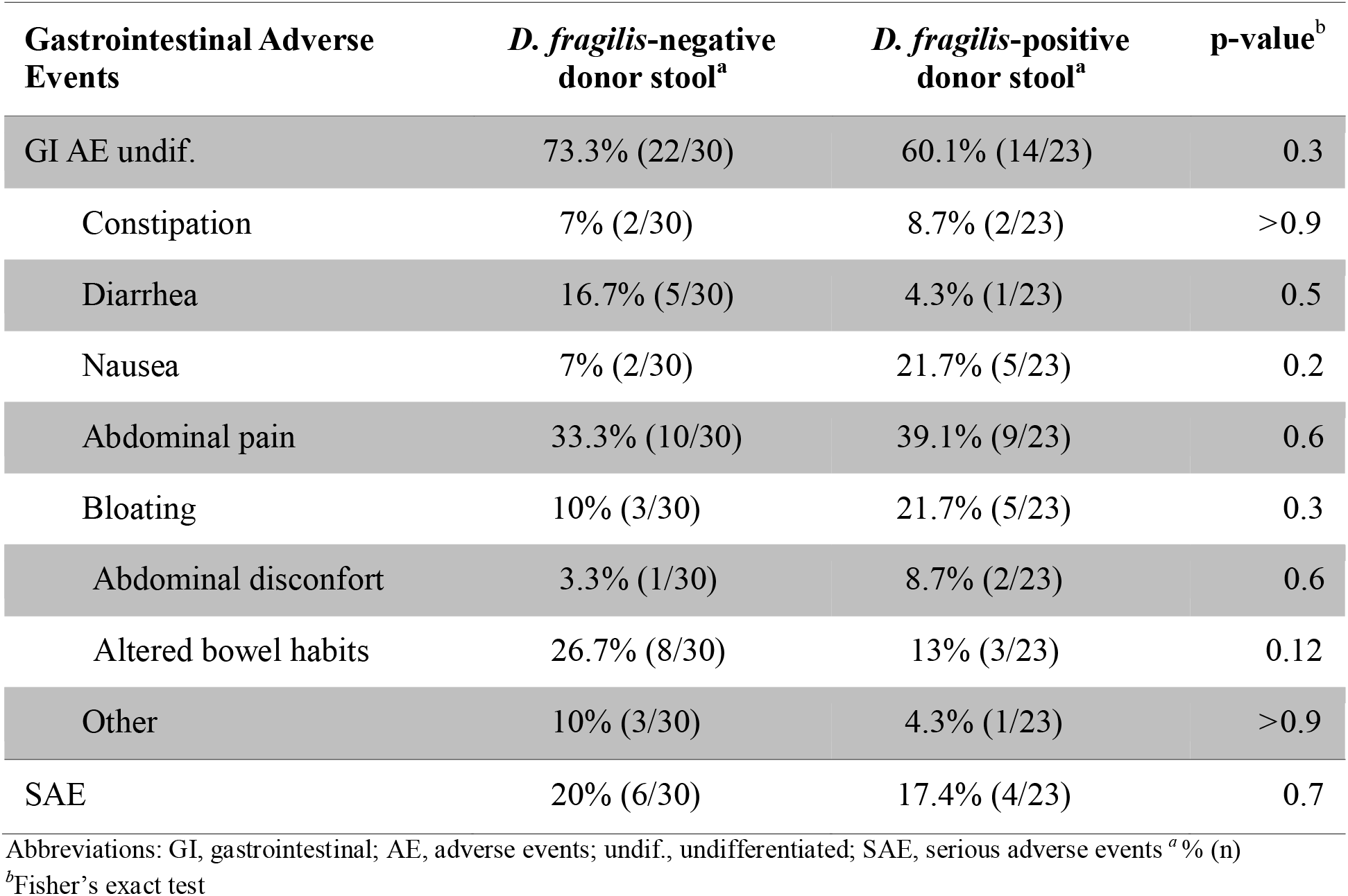
Adverse events occurring at least once during follow-up and comparison between *D. fragilis*-positive and -negative donor stool recipients.

## DISCUSSION

The framework of FMT represents an excellent opportunity to study the debated pathogenicity of the enteric protozoan *D. fragilis*. To assess the implication of *D. fragilis* colonization on gastrointestinal health, we assessed the presence of *D. fragilis* in stools from donors as well as their paired FMT recipients alongside their mid-term clinical symptoms. To our knowledge, this is the first report of *D. fragilis* detection by RT-PCR in stool samples of previously negative recipients, following FMT. Although many donors tested positive for *D. fragilis* (40%), the new detection of this protozoan in recipients post-FMT via RT-PCR was rare, occurring in only 4 out of 22 cases. Neither the safety as measured by the occurrence of AE, nor the efficacy of FMT was affected by the detection of *D. fragilis* donors within this cohort.

The life cycle of *D. fragilis* remains incompletely understood. While rare putative cyst and precyst forms have been observed in clinical specimens, the precise modes of human transmission remain unclear [9,16]. While direct fecal-oral transmission has yet to be confirmed [9], our results are suggestive of some level of transmission by this route and may support the hypothesis of the existence of a cyst stage in *D. fragilis* [17]. This finding, however, is contrary to previous studies who found no evidence of *D. fragilis* transmission from donor to recipient via FMT in rCDI patients [18]. Moreover, the lack of a reproducible animal model for *D. fragilis* infection has significantly hindered research into its biology and pathogenicity.

Given the fragility of this protozoan, its survival during FMT production is questionable. As demonstrated by Hurych *et al.*, a dramatic decrease in the viability of this protist after deep freezing is measured, with no viable organisms detectable in culture media after a single freeze-thaw cycle [19]. It is important to note that while PCR is a highly sensitive screening tool, it does not provide information on the viability or infectivity of *D. fragilis* but only detects the presence of nucleic acids. However, the persistent finding of DNA for several weeks after FMT would suggest a successful colonization. Our findings, showing that 81.8% of recipients remained RT-PCR negative after receiving a *D. fragilis*-positive FMT, along with existing evidence, suggest that *D. fragilis* only occasionally remains viable in the context of FMT. While we attempted a shotgun sequencing approach to confirm the donor to recipient transmission of *D. fragilis* via FMT, the low concentration of the protozoan in stools as well as the lack of a good reference genome for *D. fragilis* complicated the analysis. The number of *D. fragilis* sequences recovered was too low to assert the genetic similarity of donor and FMT recipient strains. While unlikely, the four recipients who tested positive may also have been colonized by another natural source within 15 days post-FMT.

However, the critical question is not merely whether transmission occurs, but what the clinical consequences for recipients may be. Indeed, the pathogenicity of *D. fragilis* remains controversial. While it has been associated with gastrointestinal symptoms such as diarrhea and abdominal pain, the evidence supporting this link is inconsistent. A randomized controlled trial in Denmark by Röser et al. evaluated metronidazole for *D. fragilis* infections in 96 children, finding no significant symptom improvement over placebo, though parasitological eradication was more frequent in the metronidazole group [20]. More recently, a case-control study found no significant association between the detection of *D. fragilis* and the development of gastrointestinal symptoms [21]. Similarly, several case-control studies have failed to establish a link between *D. fragilis* and conditions such as irritable bowel syndrome (IBS) or celiac disease, aligning with findings for *B. hominis* [22-24].

Advances in molecular detection have demonstrated that *D. fragilis* is commonly found in asymptomatic individuals, consistent with the results observed in our donor cohort. A Danish study found *D. fragilis* prevalence at 43% in adults and 63% in children, indicating its high occurrence in the general population [25]. Likewise, in our cohort, stool donors—selected based on stringent safety criteria—frequently tested positive for *D. fragilis*. These findings support the hypothesis that *D. fragilis* may be a commensal organism within the normal gut microbiota, rather than a pathogenic entity. In our cohort, the low prevalence of these microorganisms in patients with dysbiosis before FMT further supports this model (2% in recipients versus 40% in healthy donors, p<0.05), and even suggest that *D. fragilis* colonization may rather reflect gastrointestinal health.

Recruiting stool donors for FMT is inherently challenging due to stringent screening criteria. As noted, *D. fragilis* is commonly found in asymptomatic individuals and is often a reason for donor exclusion [8]. In our study, had we applied *D. fragilis* positivity as an exclusion criterion, 40% of potential donors would have been excluded, resulting in the discarding of 287 FMT treatments derived from these donors. Given the complexity of donor selection, this would have significantly compromised the feasibility of producing this treatment and would have deprived patients of valuable treatments, as all recipients of *D. fragilis*-positive FMT of this study were cured. Our data suggest that *D. fragilis* colonization in donors, comparably to what has been demonstrated for *B. hominis* by Terveer et al., did not adversely affect recipient outcomes, including in high-risk patients [12]. This further supports the reconsideration of *D. fragilis* as an exclusion criterion for FMT donors.

## CONCLUSIONS

Although *D. fragilis* has traditionally been considered pathogenic, its role in human health remained debated. FMT offered a valuable model for evaluating the pathogenicity of such protozoans with unclear or controversial clinical significance. Our findings suggest that FMT derived from *D. fragilis*-positive donor stools does not lead to gastrointestinal adverse events or severe outcomes in recipients, including those who are severely immunocompromised. These findings support reconsidering the exclusion of donors based on *D. fragilis* positivity, in line with the precedent set for *B. hominis* in the EDQM guidelines. Rationalizing donor screening processes could improve the availability of FMT without compromising safety, provided that appropriate clinical follow-up of recipients is ensured.

## Data Availability

The data that support the findings of this study are openly available in Mendeley data at https://data.mendeley.com/datasets/3f37czt42n/1 , DOI: 10.17632/3f37czt42n.1. Data supporting the findings is also available in the article's supplementary materials.

## Author contributions

T.G., N.K and C.B. conceived and designed the study. K.M., E.M.B, T.G., A.B. and M. A. collected the data and samples. M.C., B.N. and M.-T.R. performed molecular analyses such as RT-PCR. T.P. and C.B. performed the shotgun sequencing and analysis of stools. C.B., S.G., D.J., N.K., T.G., K.M. and A.B. contributed to analysis and interpretation of results. T.G. and K.M. contributed equally to manuscript writing. All coauthors reviewed the results and approved the final version of the manuscript.

## Personal acknowledgments

The authors thank Alexandra Mitouassiwou-Samba, Valérie Sormani, Fabienne Aparicio and Benoit Guery as well as the sequencing platform of the DMLP for their excellent technical support.

## Generative AI

During the preparation of this work the authors used ChatGPT (OpenAI, 2025) in order to rephrase certain sentences. After using this tool, the authors reviewed and edited the content as needed and take full responsibility for the content of the publication.

## Abbreviations used in this paper

AE: Adverse Events
AGA: American Gastroenterological Association
*B. hominis*: *Blastocystis hominis*
CDI: Clostridioides difficile Infection
CHUV: Centre Hospitalier Universitaire Vaudois
Ct: Cycle threshold
*D. fragilis*: *Dientamoeba fragilis*
EDQM: European Directorate for the Quality of Medicines & HealthCare
ESCMID: European Society of Clinical Microbiology and Infectious Diseases
FMT: Fecal Microbiota Transplantation
GvHD: Graft-versus-Host Disease
IBS: Irritable Bowel Syndrome
ICH E2A: International Conference on Harmonisation, article E2A (Clinical Safety)
ID: Identifier
rCDI: Recurrent CDI
REDCap: REDCap, Research Electronic Data Capture software
RT-PCR: Real-Time Polymerase Chain Reaction
SAE: Serious Adverse Events

---

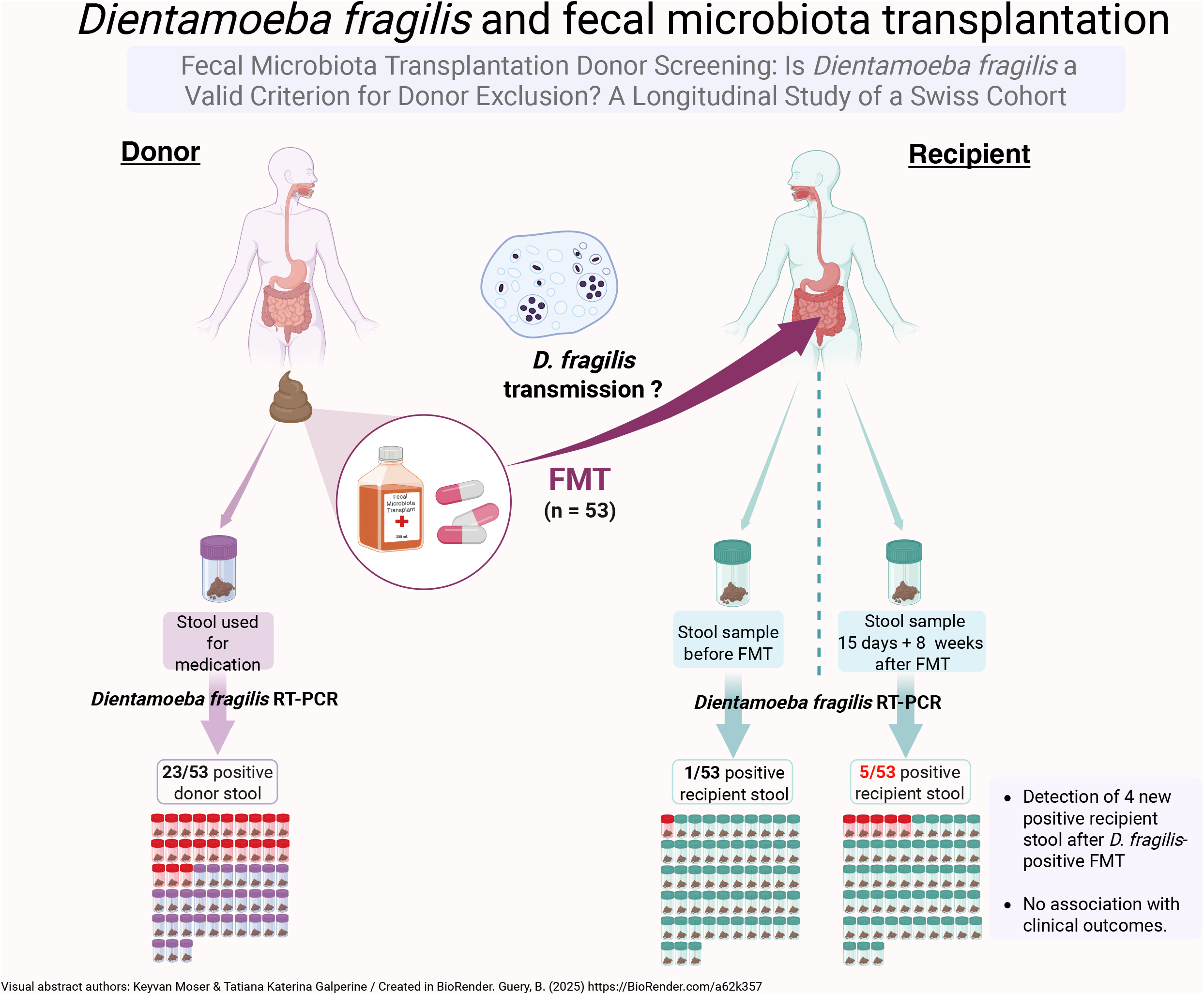

