## Supplementary material for "Fecal Microbiota Transplantation Donor Screening: Is *Dientamoeba fragilis* a Valid Criterion for Donor Exclusion? A Longitudinal Study of a Swiss Cohort": Tables 1-2-3

Table 1: Fecal microbiota transplantation with *D. fragilis*-positive donor stool and recipient *D. fragilis* status

**Donors	Recipients**

| Donor ID | *D. fragilis*  Ct value | Storage duration pre-FMT^a^, days | Patient ID | *D. fragilis* status  Pre-FMT | *D. fragilis* status  D15 Post-FMT | Ct value D15 | *D. fragilis* status  W8 Post-FMT | Ct value W8 |
| --- | --- | --- | --- | --- | --- | --- | --- | --- |
| DL01 | 23 | 237 | RL01 | Pos | Neg | n/a | Pos | 30.6 |
| DL01 | 23 | 222 | RL02 | Neg | Neg | n/a | Neg | n/a |
| DL01 | 23 | 217 | RL03 | Neg | Neg | n/a | Neg | n/a |
| DL01 | 23 | 168 | RL05 | Neg | Neg | n/a | Neg | n/a |
| DL01 | 23 | 204 | RL05 | Neg | Neg | n/a | Neg | n/a |
| DL01 | 23 | 162 | RL07 | Neg | Neg | n/a | Neg | n/a |
| DL01 | 23 | 203 | RL08 | Neg | Neg | n/a | Neg | n/a |
| DL01 | 23 | 35 | RL10 | Neg | Neg | n/a | Neg | n/a |
| DL01 | 27.2 | 239 | RL35 | Neg | Neg | n/a | Neg | n/a |
| DL01 | 23 | 257 | RL36 | Neg | Neg | n/a | Neg | n/a |
| DL01 | 23 | 384 | RL38 | Neg | Pos | 25 | Pos | 21 |
| DL01 | 19.3 | 370 | RL42 | Neg | Pos | 21 | Pos | 21 |
| DL01 | 19.3 | 377 | RL45 | Neg | Neg | n/a | Neg | n/a |
| DL01 | 23 | 729 | RL47 | Neg | Neg | n/a | Neg | n/a |
| DL02 | 23.13 | 202 | RL04 | Neg | Neg | n/a | Neg | n/a |
| DL02 | 23.13 | 246 | RL37 | Neg | Neg | n/a | Neg | n/a |
| DL02 | 23.13 | 379 | RL46 | Neg | Pos | 27 | Pos | 26 |
| DL03 | 32.5 | 260 | RL06 | Neg | Neg | n/a | Neg | n/a |
| DL03 | 31.6 | 106 | RL27 | Neg | Pos | 33.6 | Pos | 34.5 |
| DL03 | 31.6 | 462 | RL43 | Neg | Neg | n/a | Neg | n/a |
| DL03 | 31.6 | 497 | RL44 | Neg | Neg | n/a | Neg | n/a |
| DL05 | 36.1 | 714 | RL11 | Neg | Neg | n/a | Neg | n/a |
| DL05 | 30.5 | 702 | RL11 | Neg | Neg | n/a | Neg | n/a |

Abbreviations: Ct, cycle threshold; FMT, fecal microbiota transplantation; ID, identification; n/a, not applicable; Neg, negative; Pos, positive; D15,

15 days post-FMT; W8, 8 weeks post-FMT

*^a^*Time elapsed between stool donation and FMT administration

Table 2: Demographic characteristics of fecal microbiota transplantation recipients and comparison between transfer of *D. fragilis*-positive and -negative donor stool

| **Characteristics** | ***D. fragilis*-negative donor stool^a^** | ***D. fragilis*-positive donor stool^a^** | **p-value**^b^ |
| --- | --- | --- | --- |
| Age | 73 (44, 79) | 57 (52, 71) | 0.2 |
| Sex |  |  | *>*0.9 |
| Male | 73% (22/30) | 74% (17/23) |  |
| Female | 27% (8/30) | 26% (6/23) |  |
| Severe CDI | 50% (15/30) | 48% (11/23) | 0.9 |
| Immunosuppression | 27% (8/30) | 43% (10/23) | 0.2 |
| Severe immunosuppression | 6.7% (2/30) | 8.7% (2/23) | *>*0.9 |
| Route of FMT administration |  |  | 0.05 |
| Capsules | 70% (21/30) | 43% (10/23) |  |
| Colonoscopy | 30% (9/30) | 48% (11/23) |  |
| Jejunostomy | 0% (0/30) | 8.7% (2/23) |  |
| FMT efficacy |  |  | 0.12 |
| Cured | 87% (26/30) | 100% (23/23) |  |
| Recurrence | 13% (4/30) | 0% (0/23) |  |

Abbreviations: CDI, *Clostridioides difficile* infection; FMT, fecal microbiota transplantation

*^a^* Median (Q1, Q3); % (n) *^b^* Wilcoxon rank sum test; Fisher’s exact test; Pearson’s *χ*^2^ test

Table 3: Adverse events occurring at least once during follow-up and comparison between

*D. fragilis*-positive and -negative donor stool recipients

| **Gastrointestinal Adverse**  **Events** | ***D. fragilis*-negative donor stool^a^** | ***D. fragilis*-positive donor stool^a^** | **p-value**^b^ |
| --- | --- | --- | --- |
| GI AE undif. | 73.3% (22/30) | 60.1% (14/23) | 0.3 |
| Constipation | 7% (2/30) | 8.7% (2/23) | *>*0.9 |
| Diarrhea | 16.7% (5/30) | 4.3% (1/23) | 0.5 |
| Nausea | 7% (2/30) | 21.7% (5/23) | 0.2 |
| Abdominal pain | 33.3% (10/30) | 39.1% (9/23) | 0.6 |
| Bloating | 10% (3/30) | 21.7% (5/23) | 0.3 |
| Abdominal disconfort | 3.3% (1/30) | 8.7% (2/23) | 0.6 |
| Altered bowel habits | 26.7% (8/30) | 13% (3/23) | 0.12 |
| Other | 10% (3/30) | 4.3% (1/23) | *>*0.9 |
| SAE | 20% (6/30) | 17.4% (4/23) | 0.7 |

Abbreviations: GI, gastrointestinal; AE, adverse events; undif., undifferentiated; SAE, serious adverse events *^a^* % (n)

*^b^*Fisher’s exact test
