## Supplementary Table 1 for "Fecal Microbiota Transplantation Donor Screening: Is *Dientamoeba fragilis* a Valid Criterion for Donor Exclusion? A Longitudinal Study of a Swiss Cohort"

**Supplementary Table 1:** Biological screening for donor selection in fecal microbiota transplantation for the treatment of *Clostridioides difficile* infection

Blood Analysis

Hematology: complete blood count

Chemistry: Creatinine, C reactive protein, albumin, liver enzymes, glycated hemoglobin

Viruses: Hepatitis A (IgM anti-HAV), B (HBsAg, total anti-HBc, anti-HBs), Hepatitis C antibody, and hepatitis E (IgM, IgG), HIV (anti-HIV 1/2, p24 antigen), HTLV-1/2 (anti-HTLV-1/2)

Bacteria: *Treponema pallidum* antibodies (TPHA)

Parasites: Strongyloides stercoralis (serology) (only if travel to a risk area), Entamoeba histolytica (serology)

Stool Analysis:

Bacteria: Clostridioides difficile, Campylobacter jejuni/coli, Salmonella spp, Shigella spp, Entero-invasive E. coli, shiga toxins STx1 and STx2, Yersinia enterocolitica, *Helicobacter* pylori

Viruses: Adenovirus, rotavirus, norovirus groups I and II, *Hepatitis E*, SARS-CoV-2

Parasites: Giardia intestinalis, Cryptosporidium hominis/parvum, Entamoeba histolytica, helminths, Dientamoeba fragilis, Strongyloides stercoralis, Blastocystis hominis

Multi-resistant bacteria: carbapenemase-producing Enterobacterales, extended-spectrum beta-lactamases vancomycin-resistant enterococci, Methicillin-resistant *Staphylococcus aureus*

Immunological marker: Calprotectin

For severely immunosuppressed patients

- Blood: Cytomegalovirus (CMV) IgM and IgG
- Epstein-Barr virus (EBV) IgM and IgG, toxoplasmosis (IgG, IgM)
- Stool: *Plesiomonas shigelloides*, parechovirus, astrovirus, enterovirus, sapovirus, *Cyclospora*, *Isospora*, *Microsporidia*
