## Supplementary Material for "Fecal Microbiota Transplantation Donor Screening: Is *Dientamoeba fragilis* a Valid Criterion for Donor Exclusion? A Longitudinal Study of a Swiss Cohort"

**FMT production**

The stool donation campaign at the CHUV Center lasts four weeks, commencing and concluding with a thorough screening process for donors according to an ongoing Swissmedic regulations based on the current version of the European Directorate for the Quality of Medicines & HealthCare (EDQM).

Between 50 and 70 grams of feces were used for FMT production of capsules and suspensions respectively. For suspension preparation, the freshly collected donation was first diluted with 0.9% NaCl, then filtered using sterile non-woven gauze, and cryopreserved with glycerol to a final concentration of 10%. For capsule preparation, the freshly collected donation was diluted with 0.9% NaCl, filtered then centrifuged. After discarding the supernatant, the pellets were resuspended in glycerol, filtered and double-encapsulated in modified-release and ready-to-administer 0 and 00 capsules (Capsugel® DRCaps, Lonza, Switzerland).

FMT products were stored at -80°C for up to 24 months. Before use, FMT suspensions were thawed either overnight at 2-8°C or in a water bath at maximum 35°C for up to 90 minutes, then filtered and repackaged in syringes.

**Diagnosis of *D. fragilis* RT-PCR**

RT-PCR assays were performed using either native stool samples or stool samples preserved in BHIG. Preliminary tests on 10 reference samples with known presence of the pathogen demonstrated reproducible RT-PCR results regardless of the sample type.

DNA extraction: DNA was extracted using the DNA Mini kit protocol (Qiagen, 51306) with minor modifications. For native stool samples, 100 mg of fresh stool was re-suspended in 1 ml of PBS. For feces preserved in BHIG medium, 500uL of BHIG stool mixture was mixed with 1ml of Phosphate-buffered-saline (PBS), vortexed and centrifuged (13 000g for 10min) before the pellet was re-suspended in 500 uL PBS. Afterwards both were centrifuged at 14'000 rpm for 5 min. The supernatant was removed and the pellet re-suspended in 400uL of ATL buffer and 40uL of proteinase K followed by an incubation at 56°C for 2 hours interrupted by short mixing every 30 min. Later on, the samples were vortexed and 200uL of supernatant was further processed according to the kit protocol. DNA was eluted in 200uL AE buffer and used for qPCR.

qPCR amplification: The 18S rRNA *Dientamoeba fragilis* specific RT-PCR protocol was conducted using TaqMan GenExpression Master Mix (Thermo Fisher, Switzerland, 4369016). The following sense primers, antisense primers and probe were used to amplify a 109bp fragment of *D. fragilis*: (5’-to-3’ forward primer CAA CGG ATG TCT TGG CTC TT [1], 5’-to-3’ reverse primer AAT ACG CAA TGT GCA TTC AAA G [from Diagnostic center, Swiss TPH, self-designed], probe (hexachlorofluoroscein [HEX]- CAA TTC TAG CCG CTT AT-MGB-NFQ) (1) (Eurofins Genomics, Ebersberg, Germany).

The thermo profile for both qPCR on the Quantstudio 5 real time PCR machine (Thermo fischer) was 2 min at 50°C and 10 min at 95 °C followed by 40 cycles of 15s at 95°c and 1 min at 58°C. On each plate negative, internal and positive controls (plasmids with a concentration of 10^3^ containing an insert with the sequence of *D. fragilis*) real time PCR product were included. A cut-off cycle threshold (Ct) of <40 was used for both PCRs.

**Metagenomics**

For shotgun metagenomics sequencing, libraries were prepared using the Illumina DNA Prep kit before sequencing on a NextSeq 1000 (Illumina, USA), allowing to retrieve between 35.8 and 50.97 million reads per sample. Given that no genome sequence is available for *D. fragilis*, the 6,595 transcripts described by Barratt et al. [1] were used for read mapping using bowtie v2.5.4 with default parameters [2].

[1] Barratt JLN, Cao M, Stark DJ, Ellis JT. The Transcriptome Sequence of Dientamoeba fragilis Offers New Biological Insights on its Metabolism, Kinome, Degradome and Potential Mechanisms of Pathogenicity. Protist. 2015 Sep;166(4):389–408.

[2] Langmead, B., Salzberg, S. Fast gapped-read alignment with Bowtie 2. Nat Methods 9, 357–359 (2012). https://doi.org/10.1038/nmeth.1923
